# Novel Parkinson’s Disease Genetic Risk Factors Within and Across European Populations

**DOI:** 10.1101/2025.03.14.24319455

**Authors:** The Global Parkinson’s Genetics Program (GP2), Hampton L. Leonard

## Abstract

**Introduction:** We conducted a meta-analysis of Parkinson’s disease genome-wide association study summary statistics, stratified by source (clinically-recruited case-control cohorts versus population biobanks) and by general European versus European isolate ancestries. This study included 63,555 cases, 17,700 proxy cases with a family history of Parkinson’s disease, and 1,746,386 controls, making it the largest investigation of Parkinson’s disease genetic risk to date.

**Methods:** Meta-analyses were performed using standard fixed and random effect models for the European sub-populations, the case-control studies, and the population biobanks separately. Finally, all of the European ancestries for all study types as well as proxy cases were combined in our final cross-European meta-analysis. We estimated heritable risk across ancestry groups, investigated tissue and cell-type enrichment, and prioritized risk genes using public data to facilitate functional follow-up efforts.

**Results:** The final combined cross-European meta-analysis identified 134 risk loci (59 novel), with a total of 157 independent signals, significantly expanding our understanding of Parkinson’s disease risk. Multi-omic data integration revealed that expression of the nominated risk genes are highly enriched in brain tissues, particularly in neuronal and astrocyte cell types. Additionally, we prioritized 33 high-confidence genes across these 134 loci for future follow-up studies.

**Conclusions:** By integrating diverse European populations and leveraging harmonized data from the Global Parkinson’s Genetics Program (GP2), we reveal new insight into the genetic architecture of Parkinson’s disease. We identified a total of 134 risk loci, expanding the number of known loci associated with PD by approximately 24%. We also provided an initial layer of biological context to these results through follow-up analyses in an effort to facilitate follow-up studies and precision medicine efforts with the goal of advancing Parkinson’s disease research.

## INTRODUCTION

Over the past decade, Parkinson’s disease (PD) genetics research has benefited from an open-science collaborative framework, enabling robust genome-wide association study (GWAS) meta-analyses that have so far identified 102 risk loci across diverse genetic ancestries^1,2^. Building on these efforts, the Global Parkinson’s Genetics Program (GP2) has spearheaded the next generation of community-driven research in this field^3^.

PD is estimated to affect 0.5% of the global aging population, with prevalence reaching up to 2% in individuals above 65 in European populations^4^. It is the fastest-growing neurological disorder, and projections indicate that the incidence rate could double by 2030^5,6^. A key objective of GP2 is to enable researchers to conduct inclusive and diverse GWAS^7,8^. As GP2 continues to expand research across these diverse populations, larger sample sizes will allow for more in-depth exploration of population isolates within high-risk ancestry groups, as well as a more detailed analysis of the risks associated with rare genetic variants in these populations. As GP2 expands its research across diverse populations, the increasing sample sizes will enable deeper investigation of population isolates within high-risk ancestry groups and more comprehensive analysis of rare genetic variants’ risks. A significant advantage of this study is that all GP2 cohorts—comprising 22,048 cases (26.98%) in the final meta-analysis—were genotyped using the NeuroBooster array (NBA) and processed using the same pipeline^9^. The NBA integrates approximately 2 million backbone variants from the Infinium Global Diversity Array with 95,000 custom variants associated with various neurological conditions. With over 10,000 additional tagging variants, the array enables accurate imputation of more than 15 million common variants via TOPMed across diverse populations.

In this study, we investigated the role of common genetic variation within and across specific European populations on PD risk, aiming to identify risk loci and estimate genetic risk factors enriched in Icelandic, Finnish, Ashkenazi Jewish, and general European ancestries. The final cross-European meta-analysis included 63,555 cases, 17,700 proxy cases with a family history of PD, and 1,746,386 controls free of neurological diseases. We then assessed heterogeneity and heritability and leveraged public multi-omics data to prioritize genes within loci. This study highlights the importance of large, harmonized sample sizes, such as those generated by GP2, and emphasizes the value of multi-ancestry analyses—even within European populations—in advancing our understanding of disease biology by identifying novel risk loci and characterizing genetic differences across sub-populations.

## METHODS

### Study design

Our study was divided into two analysis arms. The first focused on subsets of European ancestry, as determined by custom ancestry estimation algorithms for individual level data and country of origin for population summary stats, which included general European, Icelandic, Finnish, and Ashkenazi Jewish populations, which will be abbreviated throughout the paper as EUR, ICE, FIN, and AJ, respectively^10^. The second analysis arm focused on case ascertainment criteria, including clinically diagnosed cases based on the Queen Square Brain Bank Criteria for PD or similar diagnostic criteria, electronic medical records using ICD-10 codes from biobanks, or proxy case status, similar to previous GWAS in this area^2^. The study design is illustrated in **Figure 1**. Fixed and random-effect meta-analyses were conducted for each separate ancestry group as well as for traditional case-control studies only and population/proxy case biobank cohorts only. Finally, a joint analysis combining all ancestry groups and study type was conducted, which was the primary focus of our study, as it maximized the use of all available samples.

**Figure 1:**
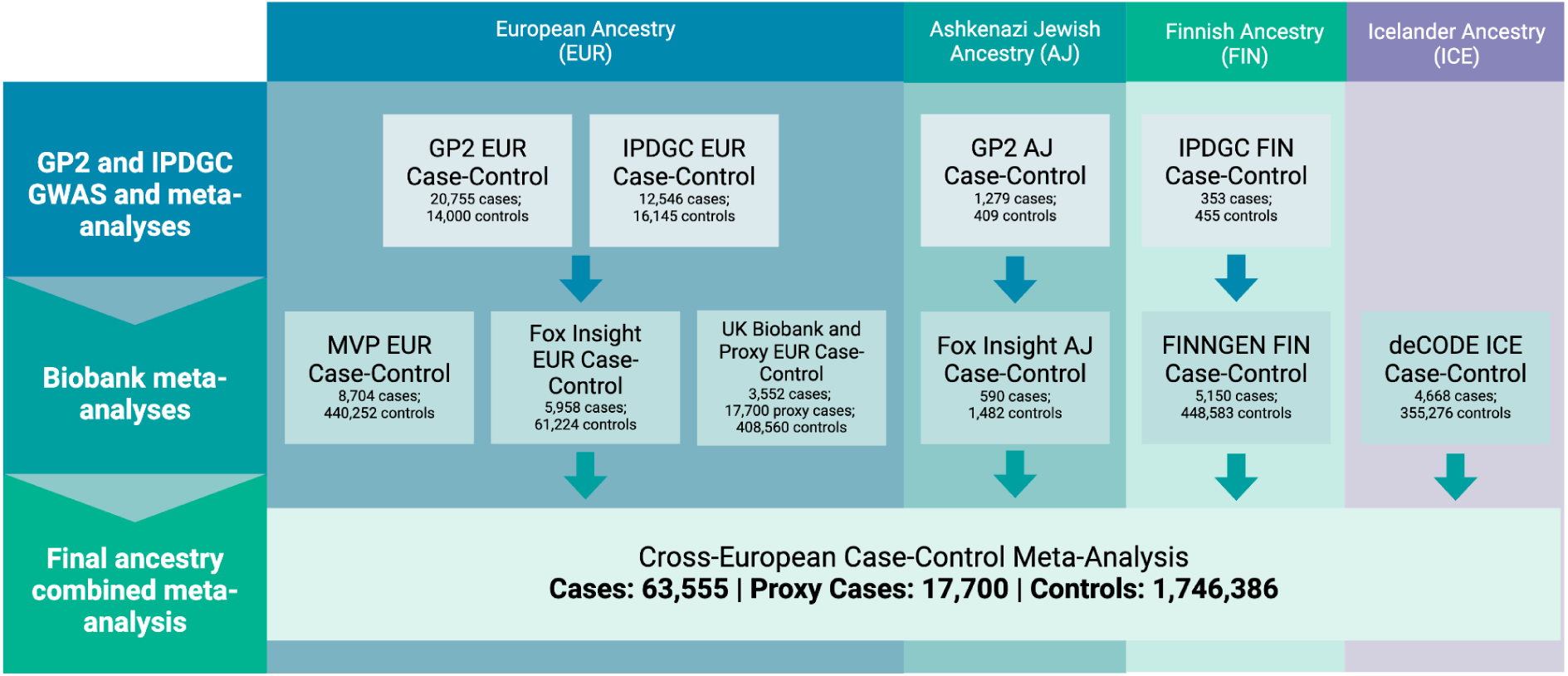
Flowchart detailing study design schema and sample counts. Study design and sample size for each ancestry and dataset. Refer to Supplementary Table 1 for a more detailed breakdown of sample size per dataset.

### Datasets analyzed

For a full inventory of all datasets analyzed, please refer to **Supplementary Table S1**. The majority of cases were sourced from GP2, along with non-overlapping samples from previous datasets curated by the International Parkinson’s Disease Genetics Consortium (IPDGC)^2,3^. These studies comprised the traditional clinical case-control study portion of this analysis, as their diagnoses were primarily sourced from official criteria as part of recruitment. Both a proxy-case GWAS, as defined previously^11^, and a case-control GWAS, as defined by ICD coding criteria, were run using biobank-scale data from the UK Biobank (UKB). Summary statistics for curated PD phenotypes were downloaded from the FinnGen biobank portal, using the G6_PARKINSON endpoint from release 11^12^. Summary statistics for clinically defined cases from the Icelandic population were provided by deCODE Genetics^13^. The data used to generate deCODE statistics was approved by the National Bioethics Committee (NBC, VSN-17-142; VSNb2017060004/03.01) following review by the Icelandic Data Protection Authority. All genotyped participants signed a broad informed consent allowing the use of their samples and data in projects at deCODE genetics approved by the NBC. Due to data sharing limitations, 23andMe PD summary statistics were not included. Instead, cases from the Fox Insight Genetics Study (FIGS), which includes a subset of the 23andMe PD samples, were matched to aged population controls with comparable Illumina arrays and processed in the same manner as the GP2 samples using GenoTools^14–16^. PD summary statistics from the Million Veteran’s Program (MVP) were downloaded from dbGAP (accession number: phs002453.v1.p1)^17^. The UKB, FinnGen, deCODE, FIGS, and MVP cohorts comprised the biobank portion of this analysis.

### Cohort quality control and ancestry

GP2, FIGS, and each participating IPDGC dataset underwent standard quality control and ancestry estimation via GenoTools, described in detail elsewhere^10^. As there is known overlap between the IPDGC studies and GP2, in addition to the standard relatedness filtering within studies performed by GenoTools, duplication and relatedness to GP2 samples was assessed using KING^18^ for each IPDGC dataset. Samples that were duplicated or related to GP2 samples on a level closer than first-degree cousins were removed from the IPDGC studies, in order to prioritize GP2 samples, which were all genotyped on the same NeuroBooster Array^9^. Only predicted ‘EUR’ samples were used from the IPDGC data, as predicted ‘AJ’ ancestry sample sizes were too small (n<100 cases or controls). To construct the combined FIGS and population control dataset, we kept FIGS cases from the largest sample set that had been genotyped on the same Illumina GSA array and merged these with population controls genotyped on the Illumina GSA array from the National Cancer Institute (NCI). NCI samples were labeled as population controls if they were both free from any neurological disease and were not taking a PD drug. After QC with GenoTools, each dataset was imputed separately by genetically predicted ancestry using the TOPMed-r3 reference panel. Imputed data was then filtered for imputation quality greater than 0.3 prior to analyses.

UKB quality control was performed previously by UKBiobank^19^, however, to keep ancestry prediction methods similar, the GenoTools ancestry prediction module was run on the UKB genotyped samples. Samples predicted ‘EUR’ were then used for the case control and the proxy analyses (sample sizes for predicted ‘FIN’ and ‘AJ’ ancestries were too small). PD cases were defined by the UKB algorithmically-defined code p42032, which includes ICD9, ICD10, and self report codes curated by UKB. Controls were sampled at 20X the case sample size from anyone currently older than 60 years of age who did not have a value for p42032, did not have any reported ICD10 codes of G20, G21, G22, G23, G24, G25, and did not report family history of PD. Proxy cases were defined as anyone who reported a mother or father with PD. Proxy controls were selected using the same criteria as used for the case-control analysis but only for samples that did not not overlap with the controls used for the UKB case-control analysis. Related individuals were removed prior to analysis.

The FinnGen, deCODE, and MVP summary statistics underwent minimal quality control filtering to adjust incorrect frequencies and some problematic variants with effect estimates of 0. Since we did not have access to individual-level data for these cohorts, ancestry labels were attributed according to country of origin for FinnGen (FIN) and deCODE statistics (ICE). MVP estimated European ancestry via their own pipeline, so we included those statistics under the ‘EUR’ label.

### GWAS and Meta-analyses

GWAS were conducted using Plink2^20^, correcting for principal components, genetically-determined sex, and age (when available). Summary statistics were filtered for minor allele frequency (MAF) > 1% specific to each study, except for the *GBA1*, *SNCA*, and *LRRK2* regions (gene ±250kb). Fixed-effect and random-effects meta-analyses were carried out per ancestry and per case ascertainment criteria (clinical case-control or biobank) as described, and then finally for the full joint analysis using Plink1.9 ^21^. For each meta-analysis, we defined loci only using variants that were filtered for quality by keeping variants that were present in at least 50% of our total studies (11 out of 21) and had an I^2^ heterogeneity estimate < 80. Genomic inflation estimates were calculated for each meta-analysis. Scaled genomic inflation was nominal and below 1.03 for all meta-analyses, all lambda and scaled lambda_1000_ estimates^22^ can be found in **Supplementary Table S2**. Significant independent loci were determined by the genome-wide significance threshold of 5e-8 and by using a linkage disequilibrium (LD) clumping threshold of r^2^ > 0.6 within 250kb windows, facilitated by Functional Mapping and Annotation (FUMA) as described elsewhere previously^1^. A locus was classified as novel if it fell outside of a region previously identified by Nalls et al., Foo et al., or Kim et al., by ±250 kb. All independent variants, known and novel, can be found in **Supplementary Table S3**.

Conditional analysis was performed with the Genome-wide Complex Trait Analysis package under default settings for multi-SNP-based conditional & joint association analysis (COJO) to identify additional independent signals at each locus^23,24^. COJO performs conditioning with summary level data and infers LD information from a reference panel, using a stepwise method to iteratively add and remove variants per the genome-wide significance threshold until no additional variants can be added or removed. COJO requires a large reference panel to more accurately ascertain independence and recommends using the largest participating cohort in your study, thus the reference panel used for this analysis consisted of the individual-level GP2 general European ancestry predicted dataset, with related individuals removed (37,916 individuals of European Ancestry). The summary level input consisted of the full joint analysis summary statistics filtered for presence in 50% of the studies and I^2^ < 80.^23,24^ Any variant passing the conditional and joint analyses at 5e-8 that was not initially nominated as independent by FUMA was added to **Supplementary Table S3** as an independent signal. Some lead variants nominated by FUMA did not pass the COJO conditioning independence threshold of 5e-8, this has been annotated in **Supplementary Table S3**.

### Genetic correlation and heritability

Genetic correlations and heritability were calculated using summary statistics with linkage disequilibrium score regression (LDSC) under default settings^25^. We investigated correlation between ancestry-specific meta-analyses and between biobank and case-control meta-analyses. Proxy cases were not included in these calculations to avoid confounding heritability estimates.

### Prioritization of tissues, cell types, and genes under peaks

We used FUMA combined with annotations from Ensembl Variant Effect Predictor (VEP) to prioritize genes for future follow-up ^26,27^.

Gene-based tests from FUMA’s Multi-marker Analysis of GenoMic Annotation (MAGMA) were utilized under default settings to collapse SNVs to proximal genes and conduct tissue enrichment analysis ^28^. Tissue and cell type enrichments were carried out using FUMA, testing for enrichment of GWAS loci among genes expressed in various tissue and cell type reference datasets from mice and humans. For gene prioritization under peaks, we implemented a Polygenic Priority Score (PoPS) using the recommended defaults to leverage multi-omic data resources to identify potential functional genes for follow-up ^29^. All enrichment and prioritization analyses were based on the fixed-effect meta-analysis of all available samples to maximize statistical power.

## RESULTS

### Identification of novel and independent loci across European populations

To increase our power to nominate loci, we focused on the final joint meta-analysis including all samples of all European populations and study types. In this meta-analysis, we identified 134 genome-wide significant loci, of which 59 were more than ±250 kb from previously nominated loci. All 134 nominated loci are represented in the Manhattan plot in **Figure 2**. Genetic correlation (rG) supported our focus on the joint meta-analysis between the case-control meta-analysis and the biobank meta-analysis (LDSC rG = 0.89, SE = 0.037) (**Supplementary Table S4**). However, in order to compare results more directly across study types, association results for the case-control-only meta-analysis and the biobank-only meta-analysis are shown for all variants in **Supplementary Table S3.** A summary of novel loci identified in the final joint meta-analysis phase is presented in **Table 1**. Ancestry-specific frequencies and effects are reported in **Supplementary Table S4.** Additionally, forest plots and local association plots for all loci have been included in the **Supplementary Appendix**.

**Figure 2:**
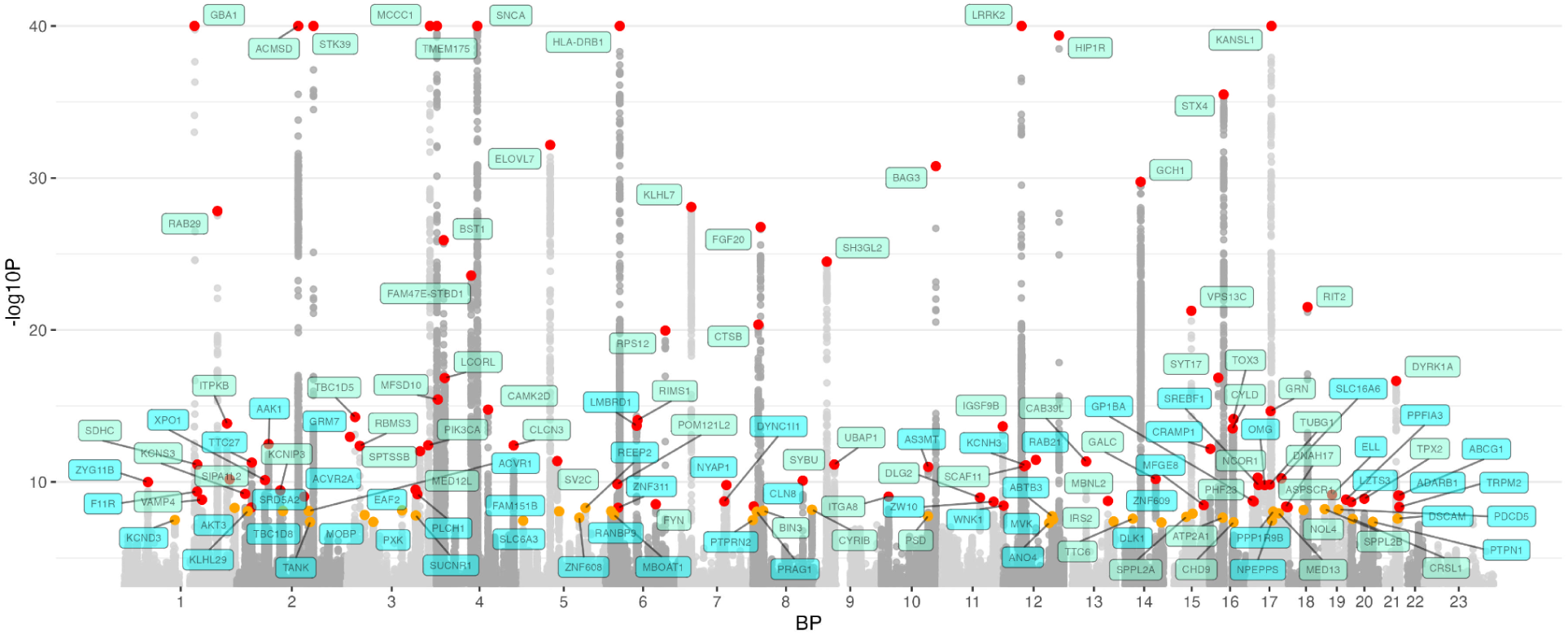
Manhattan plot of meta-analysis including all available samples. The nearest gene to each of the 134 significant loci is labeled in green for previously-identified loci and in blue for novel loci. −log10 P values were capped at 40. Variant points are color-coded, orange indicates significance levels between P < 5E-8 and P = 5E-9, while red indicates variants with P < 5E-9.

Conditional analyses carried out with COJO identified an additional 23 variants as independent signals, for a total of 157 independent variants nominated for PD risk from the combined meta-analysis and conditional analyses. Some of the independent signals nominated in previous work were replicated as independent variants here, but COJO also revealed additional independent variants in this large study, such as 3 independent coding signals in the *GBA1* locus and 3 signals in the *TMEM175* locus where previously only 2 had been nominated. New independent signals also included an additional signal in the *F11R* (rs72712893), *SDHC* (rs61802091), *STK39* (rs12618884), *TBC1D5* (rs144678625), *SUCNR1* (rs1719750116), *CLCN3* (rs1719750116), *POM121L2* (rs4713072), *BAG3* (rs1174256174), *PHF23* (rs72827590), and *BRIP1* (rs11871753) loci, and 3 additional signals in the *LRRK2* locus (rs35303786, rs17443099, rs28370650). Summary information for nominated independent signals can be found in **Supplementary Table S3.**

### Replication in previous GWAS studies

Over the past decade, previous GWAS efforts have identified 92 loci containing one or more independent genome-wide significant risk factors across general European, East Asian, and multi-ancestry meta-analyses. Here, we replicated 66 out of 78 loci from Nalls et al. (84.6%), 10 out of 11 loci from Foo et al (90.9%) (including 1 out of the 2 reported novel loci), and 65 out of 78 loci from Kim et al. (83.3%) (including 7 out of the 12 reported novel loci) at genome-wide levels of significance (within +/- 250 kb from the reported representative SNP for that locus in the previous publications).

It is worth noting that most previously significant loci that did not replicate in this study were still identified with some level of significance (75 loci at 5E-6 and 83 loci at 5E-4) but just did not meet the 5E-8 threshold. A comparison of summary information from the final joint-meta analysis and the clinical case-control meta-analysis for all 90 variants nominated in Nalls et al can be found in **Supplementary Table S5**. Additionally, a beta-beta comparison plot between the final joint meta-analysis and Nalls et al was generated for the 90 risk variants, showing high correlation (r = 0.97, P = 3.94E-52) between effects for these variants despite not all variants reaching the significance threshold in this analysis (**Supplementary Figure S1**).

### Variant associations and heritability are variable across isolates and diagnostic criteria

Significant loci in each of the FIN, ICE, and AJ analyses are summarized in **Table 2**. One potentially novel locus in Icelandic samples was detected near the gene *PTTG* (rs10031495, effect allele = T, beta = −0.1972, SE = 0.0347, p-value from GWAS = 1.32E-08), but did not replicate in the final joint-meta analysis (beta = −0.0162, SE = 0.0098, p-value = 0.1013) or meta-analyses in this study. From the joint meta-analysis of all available samples, 38 loci exhibited an index of heterogeneity (I2) greater than 30%. The heterogeneity of effect estimates across populations is graphically summarized in **Figure 3**.

**Figure 3:**
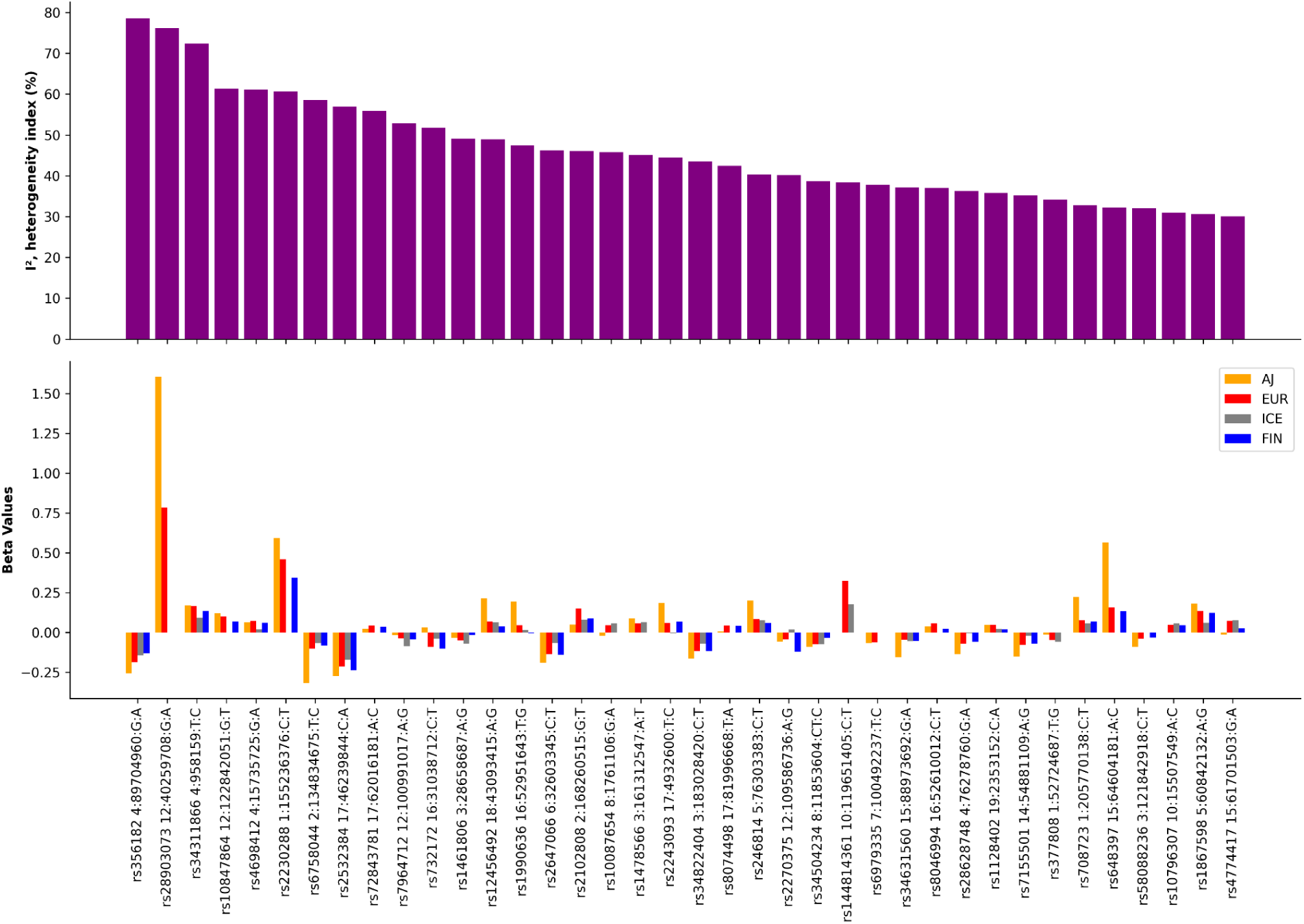
Summary of effect heterogeneity across all European ancestry population subsets. This plot focuses on loci at joint meta-analysis with I2 greater than 30% only, comparing beta estimates in AJ, EUR, ICE and FIN populations per locus as they contribute to overall heterogeneity of effect.

LDSC was used to estimate genetic correlations (rG) between the datasets included in the meta-analyses excluding proxy cases, as well as to calculate heritability. As a positive control, the rG between the GP2 EUR dataset and the non-overlapping clinically diagnosed IPDGC EUR dataset was 0.9866 (SE = 0.0684, P = 3.34E-47), confirming that we could successfully use the older IPDGC samples in the analysis with the newer, more harmonized GP2 samples. Across datasets, rG values ranged from 0.4058 to 0.9988, with the lowest observed between EUR and AJ meta-analyses (rG = 0.4058, P = 0.0048) and the highest between the EUR meta-analysis and Icelandic GWAS (rG = 0.9988, P = 0.0184). The EUR and FIN meta-analyses also showed a strong correlation, with an rG of 0.8672 (SE = 0.0726, P = 6.63E-33). When comparing clinical cases and controls with biobank-derived cases and controls within EUR datasets, the rG was 0.892 (SE = 0.0372, P = 5.64E-127). It’s important to mention that LDSC h2 was calculated using a European ancestry reference panel, which may have limited applicability to populations such as AJ, FIN, and ICE as those ancestries are not well-represented in the reference panel. While correlation was generally lower when comparing to biobank cohorts, correlation was still strong enough that we chose to focus on the results from the final combined meta-analysis. Genetic correlations across summary statistics are summarized in **Supplementary Table S6**.

As expected, heritability (h2) was lower in the biobank cohorts than in the clinical case-control studies, indicating the potential influence of differences in diagnostic sources on these estimates. We used computed liability-scaled heritability for comparisons, to account for potential biases introduced by a larger prevalence observed in case-control datasets than the general populations^30^. Interestingly, the AJ ancestry meta-analysis had the highest scaled-liability h2 estimate at 28.13%, which may reflect the very strong *LRRK2* signal seen in the AJ meta-analysis. However, the standard error was large, at SE = 25.08 scaled-liability h2 SE = 16.44 (compared to the much smaller estimate for EUR meta-analysis: scaled-liability h2 = 8.5%, SE = 0.0062), which may also explain the lower correlation seen between the EUR and AJ meta-analyses. This wide variability likely reflects the reduced precision due to a smaller case sample size and less available controls for this population and would require more follow-up. FIN and ICE datasets displayed slightly lower liability-scaled h2 estimates, 7.67% and 1.54% respectively. This can possibly be attributed to the fact that rare variants that are not included in our LDSC calculations explain more of the heritability in other populations, such as the ICE ancestry, where rare variants in *LRRK2*, *GBA1*, and *ITSN1* may explain more of the disease risk heritability in that population^13^. Observed and liability-scaled heritability estimates across summary statistics are shown in **Supplementary Table S7**.

### Functional mapping identifies genes underlying association peaks

Data from fourteen brain tissues showed significant enrichment of GWAS-associated genes in gene expression datasets analyzed using FUMA, after correcting for multiple testing. The cerebellum and frontal cortex were heavily enriched for risk genes, with the substantia nigra and spinal cord also showing significant enrichment. No significant enrichment was detected in tissues outside the brain (**Figure 4, Supplementary Table S8)**.

**Figure 4:**
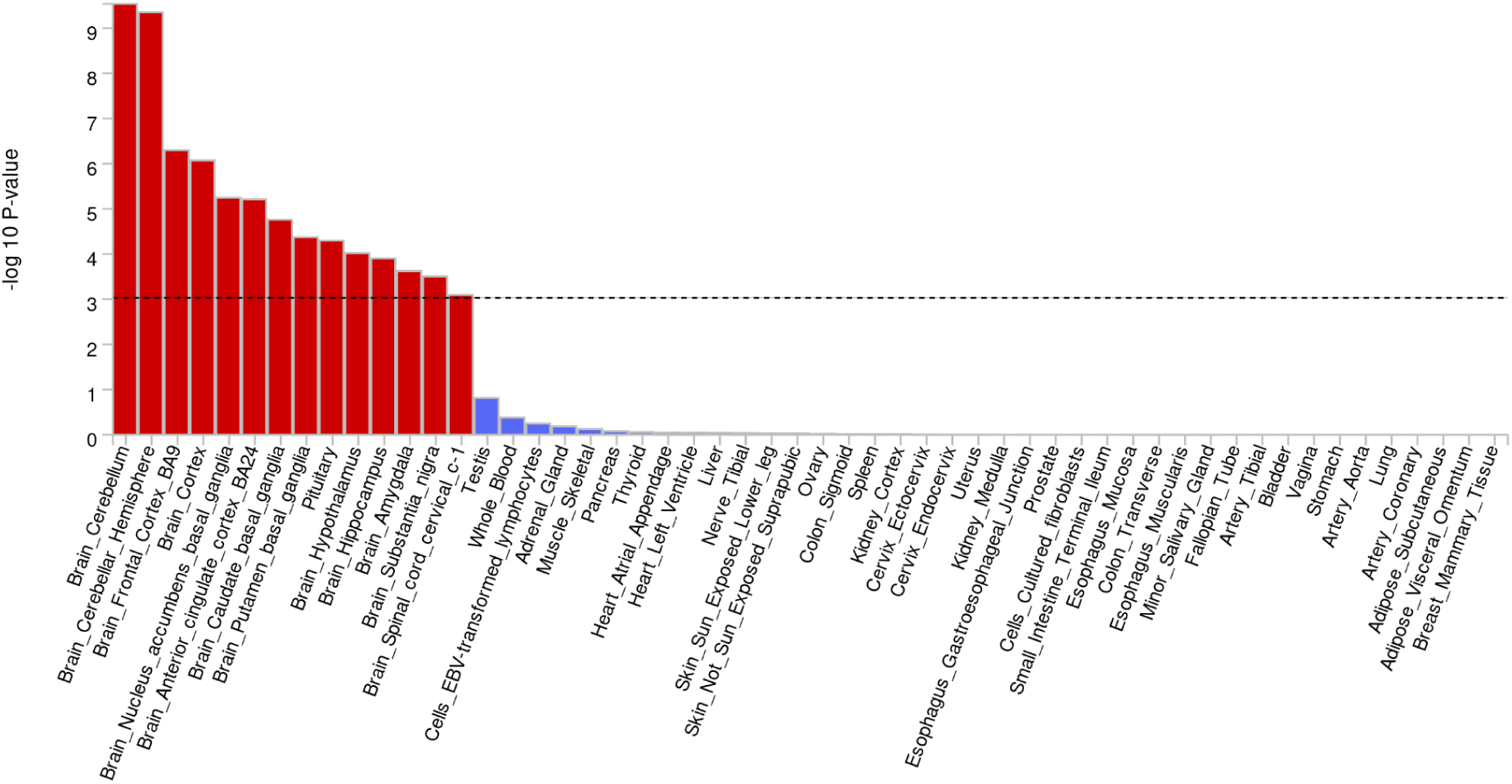
Tissue enrichments from the joint meta-analysis of all datasets suggest Parkinson’s disease genetic risk is almost exclusively relegated to the brain. Red bars denote tissues passing multiple test corrections in functional enrichment analyses.

FUMA was also used to test for enrichment across various brain-derived cell-type datasets from human and mouse sources. Neurons emerged as the predominant cell type in this enrichment analysis and TH-positive neurons showed co-enrichment with substantia nigra neurons in the same samples. Additionally, human prefrontal cortex samples revealed an enrichment of risk signals in astrocytes, which co-occurred with GABAergic neurons (**Table 3**).

PoPS identified a total of 32 genes as having a high likelihood of functional relevance based on GWAS association peaks (**Table 4**). These genes were selected using stringent criteria: passing Bonferroni correction for MAGMA gene-based tests and achieving a PoPS score in the 90th percentile or higher. Comprehensive summary statistics for all genes with a positive PoPS score, regardless of other metrics, are provided in **Supplementary Table S9**.

## DISCUSSION

This study adopted an ancestry-specific approach within the general European continental population, utilizing harmonized data from GP2 and focusing on distinct European ancestries to refine associations for improved discovery. As sample size and statistical power grow, GWAS and meta-analyses will continue to uncover rare variation and variants with smaller effect sizes, significantly broadening our understanding of the disease. In this study, we have nominated 59 novel loci associated with PD risk.

While the locus previously identified in the Icelandic population has not been replicated, it may be of interest from a biological perspective. *TBC1D1* is a Rab GTPase-activating protein involved in glucose transport, and the *TBC1D1* protein has been implicated in interactions with genes from the *VPS13* family^31^. The VPS13 protein family members, VPS13A through VPS13D, are involved in lipid transport across membranes. Each member of the VPS13 protein family has been linked to different neurological disorders^32^. Mutations in VPS13C have been previously associated with autosomal recessive early-onset PD and loss of VPS13C in human dopaminergic neurons compromises the Rab10-mediated lysosomal stress response and disrupts lysosomal structure, movement, and enzymatic activity^33,34^.

Among the genome-wide significant loci that we have replicated and discovered, to aid in the prioritization of follow-up experiments for further target due diligence, we have ranked and prioritized all potentially associated genes near nominated loci for the community by integrating multi-omics reference data. This includes a focus on 32 high-confidence genes. As well as prioritizing genes under peaks, we have also tried to alleviate the burden on target due diligence by nominating tissues, cell types, and developmental stages most likely enriched for PD risk. This should empower precision medicine as future research can more easily prioritize chasing the right gene(s), in the right tissue(s), modeling the right cell type(s) from the right sample(s) at the right stage(s) of life history.

Here, we have also taken a fine-grained approach to heritability estimates and genetic correlations across populations and datasets that were previously lumped together. We noted that 28.79% of loci exhibited heterogeneity (I^2^) greater than 30% across our European ancestry strata at the joint meta-analysis of all data, compared to Nalls et al 2019 where only 15.6% of loci had an I^2^ greater than 30%. Heritability estimates were also somewhat heterogeneous, with AJ heritability estimates at nearly double that of general European ancestry estimates, with large effect sizes in the *LRRK2* locus compared to substantially smaller but still large effect estimates in other European ancestry datasets and previously published estimates, with a roughly 2 fold increase in effect size for *LRRK2* compared to other populations. We also quantified the differences in heritability based on the source of data, with large variability in heritability estimates relating to clinic versus biobank ascertainment of cases as well as case sample size per dataset as other confounding factors. Importantly, it should be noted that the method used for estimating heritability (LDSC), can be unstable with low sample sizes, exhibiting bias and attenuated heritability estimates. Furthermore, when LDSC results include a 95% confidence interval that encompasses zero, it suggests a lack of statistical significance, which may indicate an unreliable estimate.

In this study, we used a random effects model to accommodate for the potential variations in effect sizes across studies and populations. Under the null hypothesis of no association, random effects models incorporate heterogeneity assumptions that can lead to a conservative estimation of effect sizes, potentially obscuring statistically significant findings ^35^. Particularly in multi-ancestry meta-analyses, the observed heterogeneity accounted for by random effects models may predominantly stem from differences between specific ancestral groups. For instance, the multi-ancestry meta-analysis conducted by Kim and colleagues on PD demonstrated that novel loci identified via an random effects model exhibited significant ancestral heterogeneity, with variant effects varying markedly across ancestral groups^1,2^. Similarly, an analysis of Alzheimer’s disease (AD) revealed that the heterogeneity at certain risk loci could be attributed to genetic ancestry ^36^. Consequently, the failure of random effects models to detect significant results may reflect the model’s sensitivity to ancestry-specific effects, which may not be generalizable across all studied populations^1,2^. Loci nominated in this study that pass the significance threshold with a fixed-effect model but do not pass with a random-effects model may indicate an effect that is less generalizable across different ancestries and should be evaluated in follow-up work to further investigate population-specific effects.

The limitations of this study include the use of samples of general European ancestry, as upcoming studies led by GP2 collaborators will be more inclusive as datasets grow in terms of both size and representation. For most bioinformatics software, larger and more variable linkage references as well as diverse and larger multi-omic references would be of high utility. One of the strengths of this study is the uniform processing of samples using a single array and codebase, which may have further empowered our discovery efforts. Conversely, some samples could not be re-genotyped or resequenced due to logistics or unavailability, which may introduce some noise to the analyses although this report showcases the power of data harmonization at scale.

In summary, we provide here the largest PD GWAS to date. We identified 134 loci, of which 59 are novel. There is still much to be done to follow-up and characterize these results, but we believe this work will support the next phase of PD genetic research.

## Supporting information

GP2 European GWAS Manuscript Tables

Supplementary GP2 European GWAS Manuscript Tables

## Data Availability

Summary statistics from every ancestry-level meta-analysis is available on NDPK. Code will be made available on the GP2 Github after review from the GP2 DCD.

https://ndkp.hugeamp.org/research.html?pageid=a2f_downloads_280

## ACKNOWLEDGEMENTS

This project was supported by the Global Parkinson’s Genetics Program (GP2; https://gp2.org). GP2 is funded by the Aligning Science Across Parkinson’s (ASAP) initiative and implemented by The Michael J. Fox Foundation for Parkinson’s Research (MJFF). For a complete list of GP2 members see doi.org/10.5281/zenodo.7904831. Data used in the preparation of this article were obtained from GP2. Specifically, we used Tier 2 data from GP2 release 9 (doi.org/10.5281/zenodo.14510099). Tier 1 data can be accessed by completing a form on the Accelerating Medicines Partnership in Parkinson’s Disease (AMP®-PD) website (https://amp-pd.org/register-for-amp-pd). Tier 2 data access requires approval and a Data Use Agreement signed by your institution.

This research was supported by the Aligning Science Across Parkinson’s Initiative, the Intramural Research Program, National Institute on Aging, National Institutes of Health, Department of Health and Human Services, project ZO1 AG000949, and the Michael J. Fox Foundation for Parkinson’s Research. This work utilized the computational resources of the NIH STRIDES Initiative (https://cloud.nih.gov) through the Other Transaction agreement - Azure: OT2OD032100, Google Cloud Platform: OT2OD027060, Amazon Web Services: OT2OD027852. This work utilized the computational resources of the NIH HPC Biowulf cluster (https://hpc.nih.gov).

This research has been conducted using the UK Biobank Resource under Application Number 33601.

We want to acknowledge the participants and investigators of the FinnGen study and thank them for their hard work and generosity.

We thank the participants and scientists in this study for their valuable contribution to research contributed by the deCODE genetics/Amgen Inc. team from Reykjavik, Iceland. We thank all investigators and colleagues who contributed to data collection, phenotypic characterization of clinical samples, genotyping, and analysis of the whole-genome association data associated with the deCODE data.

## DATA AND CODE AVAILABILITY

All code generated for this article, and the identifiers for all software programs and packages used, are available on GitHub (https://github.com/GP2code/GP2-EUR-metaGWAS) and were given a persistent identifier via Zenodo (DOI: 10.5281/zenodo.15013321).

Summary statistics from every ancestry-level meta-analysis are available on NDPK (https://ndkp.hugeamp.org/research.html?pageid=a2f_downloads_280).

## CONTRIBUTIONS

Hampton L. Leonard, Mary B. Makarious, Dan Vitale, and Mike A. Nalls designed the analysis, analyzed the data, and drafted the initial manuscript.

Astros Th. Skuladottir, Vala Palmadottir, Hreinn Stefansson, and Kari Stefansson designed the analysis and analyzed the data for the Iceland (ICE) ancestry GWAS from deCODE.

Kristin Levine, Hampton L. Leonard, Mary B. Makarious, Dan Vitale, Mat Koretsky contributed to quality control of GP2 genetic data and making it available to researchers.

Cornelis Blauwendraat and Andrew B. Singleton led study design, data logistics, and funding of the study.

Huw Morris, Manuela Tan, Hirotaka Iwaki, Simona Jasaityte, Ellie Stafford, Lietsel Jones, Shannon Ballard, J Solle, and Claire Wegel (Complex and Compliance Working Groups) designed the GP2 complex study, clinical and genetic data generation and preparation, and legal / data sharing logistics.

All other members of GP2 (contributors) contributed data and made critical revisions to this article.

## SUPPLEMENTARY MATERIALS

**Figure S1:**
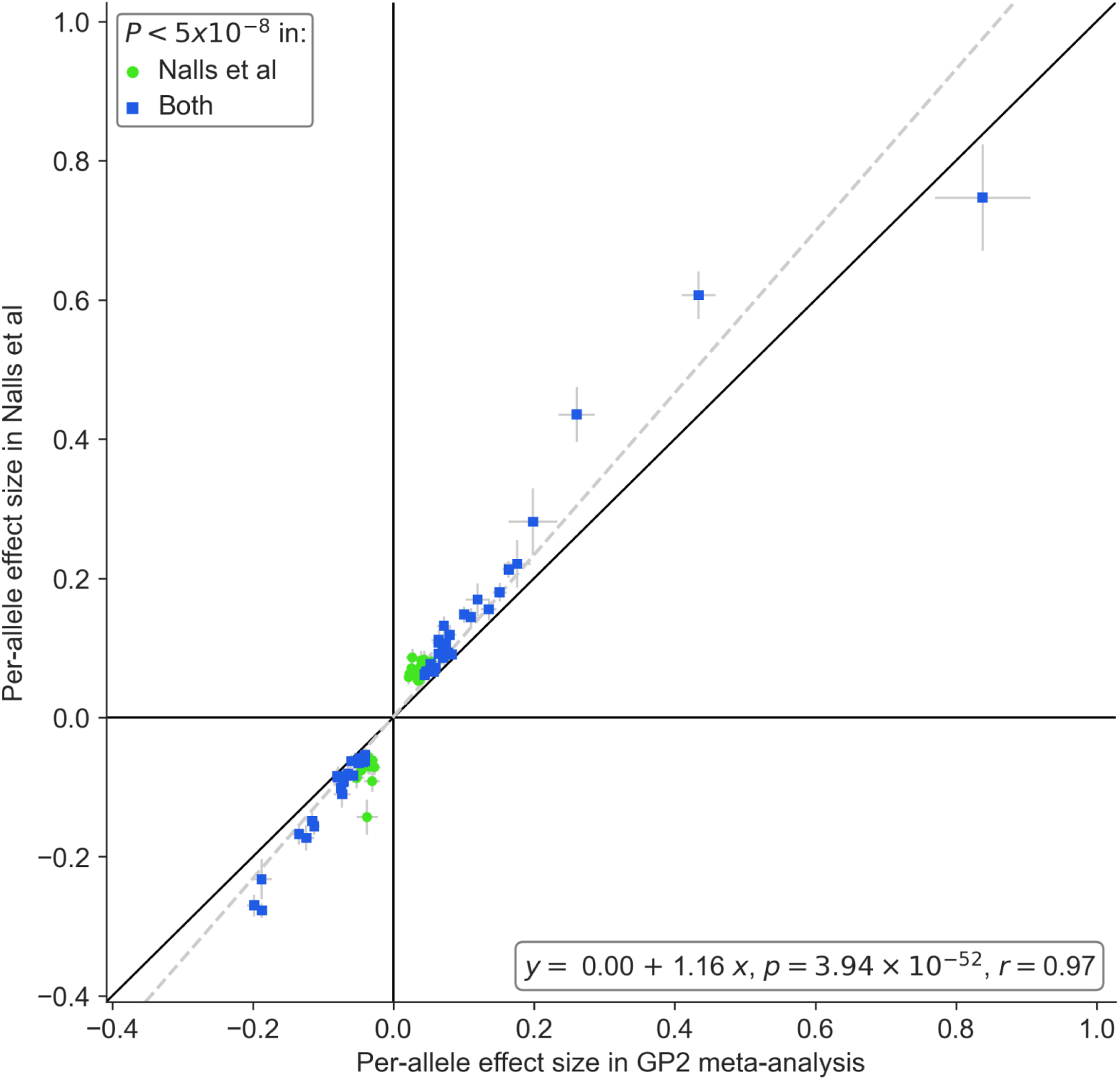
Beta-beta plot comparing effects from Nalls et al 2019 and the new joint 2025 meta-analysis for the 90 lead SNPs from Nalls et al 2019

**Supplementary Tables**

**Appendix**: Local association plots and forest plots for all loci.

